# Proton Free Induction Decay MRSI at 7T in the Human Brain Using an Egg-Shaped Modified Rosette K-Space Trajectory

**DOI:** 10.1101/2024.03.26.24304840

**Authors:** Simon Blömer, Lukas Hingerl, Małgorzata Marjańska, Wolfgang Bogner, Stanislav Motyka, Gilbert Hangel, Antoine Klauser, Ovidiu C Andronesi, Bernhard Strasser

**Affiliations:** German Center for Neurodegenerative Diseases (DZNE), Bonn, Germany; High Field MR Centre, Department of Biomedical Imaging and Image-guided Therapy, Medical University Vienna, Vienna, Austria; Center for Magnetic Resonance Research, Department of Radiology, University of Minnesota, Minneapolis, MN, USA; Christian Doppler Laboratory for Clinical Molecular MR Imaging, Medical University Vienna, Vienna, Austria; Department of Neurosurgery, Medical University of Vienna, Vienna, Austria; Advanced Clinical Imaging Technology, Siemens Healthcare AG, Lausanne, Switzerland; A. A. Martinos Center for Biomedical Imaging, Department of Radiology, Massachusetts General Hospital, Charlestown, Massachusetts, USA; Harvard Medical School, Boston, Massachusetts, USA

**Keywords:** magnetic resonance spectroscopic imaging, SNR efficiency, gradient hardware restrictions, 7 T, non-Cartesian trajectory, modified rosette trajectory

## Abstract

1.

**Purpose:** Proton (^1^H)-MRSI via spatial-spectral encoding poses high demands on gradient hardware at ultra-high fields and high-resolutions. Rosette trajectories help alleviate these problems, but at reduced SNR-efficiency due to their k-space densities not matching any desired k-space filter. We propose modified rosette trajectories, which more closely match a Hamming filter, and thereby improve SNR performance while still staying within gradient hardware limitations and without prolonging scan time.

**Methods:** Analytical and synthetic simulations were validated with phantom and in vivo measurements at 7 T. The rosette and modified rosette trajectories were measured in five healthy volunteers in six minutes in a 2D slice in the brain. A 3D sequence was measured in one volunteer within 19 minutes. The SNR, linewidth, CRLBs, lipid contamination and data quality of the proposed modified rosette trajectory were compared to the rosette trajectory.

**Results:** Using the modified rosette trajectories, an improved k-space weighting function was achieved resulting in an increase of up to 12% in SNR compared to rosette’s dependent on the two additional trajectory parameters. Similar results were achieved for the theoretical SNR calculation based on k-space densities, as well as when using the pseudo-replica method for simulated, in-vivo and phantom data. The CRLBs improved slightly, but non-significantly for the modified rosette trajectories, while the linewidths and lipid contamination remained similar.

**Conclusion:** By improving the rosette trajectory’s shape, modified rosette trajectories achieved higher SNR at the same scan time and data quality.

## 2. Introduction

Proton (^1^H)-MR spectroscopic imaging (MRSI) allows for non-invasive imaging of concentration distributions of the major metabolites in the human brain for (bio)medical applications^1,2^. MRSI studies at ultra-high field (i.e., ≥7 T) benefit from increased spectral and spatial resolution, as well as an increase in the signal-to-noise ratio (SNR)^3–6^. The most common encoding strategy for MRSI is Cartesian phase encoding. However, this approach suffers from long acquisition times^6–8^ because sampling only along the time-dimension for each k-space point is slow. In contrast, sampling along several k-space dimensions and the time-dimension simultaneously via spatial-spectral encoding (SSE) accelerates data acquisition^2,9^. Several SSE techniques have been proposed such as echo-planar spectroscopic imaging (EPSI)^10,11^, spiral-based trajectories^12,13^, rosette spectroscopic imaging (RSI)^14–16^ and concentric ring trajectories (CRT)^17–19^. Of these, self-rewinding trajectories such as rosettes and CRT benefit from higher SNR-efficiency at high spatial resolutions and spectral bandwidths due to not needing a rewinding gradient, and thus having no acquisition dead time during each circumnavigation of the trajectory. However, they need pre- and rewinder gradient at the beginning and end of all circumnavigations, thus limiting the minimum TE and TR. Hingerl et al.^19^ showed that using a CRT readout at 7 T, eight major neurometabolites could be mapped with high spatial resolution over the majority of the brain with attractive acquisition times of 3-15 minutes. However, for even higher spatial resolutions or spectral bandwidths, CRT exceed the limitations of current gradient systems^20,21^. Rosette trajectories are self-rewinding, non-Cartesian alternatives with lower hardware requirements^14^, typically implemented via rosette petals that are distributed around the k-space center. This reduces the maximum petal radius and hence the required gradient slew rate by half compared to CRT^14^. The reduced gradient stress makes rosette trajectories suitable for application at ultra-high fields and very high spatial resolutions. However, this comes at the cost of an increased acquisition time. To overcome this compressed sensing (CS) techniques have been implemented together with a 3D modified rosette trajectory, benefitting from its incoherent sampling pattern^22–24^. Compared to density-weighted-CRT^18^ the rosette trajectory has a reduced SNR efficiency due to unfavorable k-space weighting. Kasper et al. showed that SNR can be maximized if no k-space density compensation needs to be performed because the k-space weighting function of a trajectory already matches the desired target weighting (density-weighting)^25^. In addition to this SNR improvement, no density compensation during reconstruction is necessary^26^. Since ^1^H-MRSI is often strongly affected by lipid and other nuisance signals, a Hamming weighted k-space is a desirable density function to reduce signal leakage^18^. Rosette trajectories deviate substantially from the desired Hamming k-space weighting.

Thus, we propose an egg-shaped modified rosette trajectory that allows for a more SNR-efficient sampling of k-space by changing the shape of the rosette petals. Without any increase in acquisition time and with an adjustable demand on the gradient system, modified rosette trajectories present an enhancement to rosettes and are well-suited for the use at ultra-high field strengths and high spatial resolutions.

## 3. Method

### 3.1 Trajectory design

In contrast to the rosette trajectory that samples the entire k-space along a continuous trajectory, our modified rosette trajectory consists of *n* individual petals in k-space with radius *k_max_*/2, and a time varying compression factor *e(t).* It can be written as:

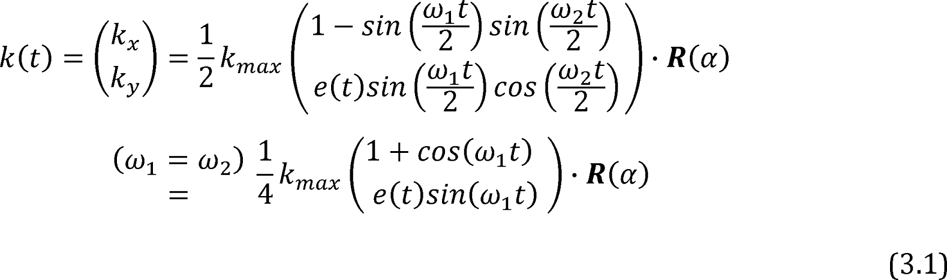

where 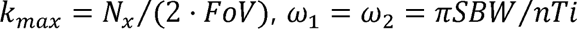 with ω_1 and_ ω_2_ being frequencies of oscillation along the radial and angular directions, respectively, *nTi* is the number of temporal interleaves, and *SBW* the spectral bandwidth. Please note that for the modified rosette trajectory ω_1 and_ ω_2_ must be equal. The factors ½ and ¼ are necessary to cover the whole k-space extent from –k_max_/2 to +k_max_/2 when rotating the trajectory with the rotation matrix *R(α).* The time varying compression factor is described by

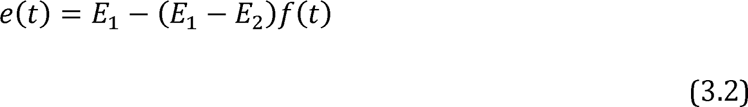

where *E*_1,_ *E*_1,_ are the boundary factors of the compression. *f(t)* is a function with a range of 0 ≤ *f(t) ≤ t* for 0 ≤ *t ≤ T,* where *T* is the time it takes to traverse each individual petal, so that *f* (*t* = 0) = *f* (*t* = *T*) = 0 and *f* (*t* = *T*/2) = 1. Here *f* (*t*) was chosen as *f(t) = sin (ξ · t/2),* which results in petals with the boundary compression factors *E*_1_ influencing the compression in the k-space center and *E*_2_ at the position *k(T/2)* = *k_max · ξ_* is the modulation frequency of the boundary compression factors and is equal to ω fulfill the above conditions.

To acquire the entire k-space, each individual petal is rotated around the k-space center with varying angle α using the rotation matrix *R(α)* (Figure 1).

**Figure 1:**
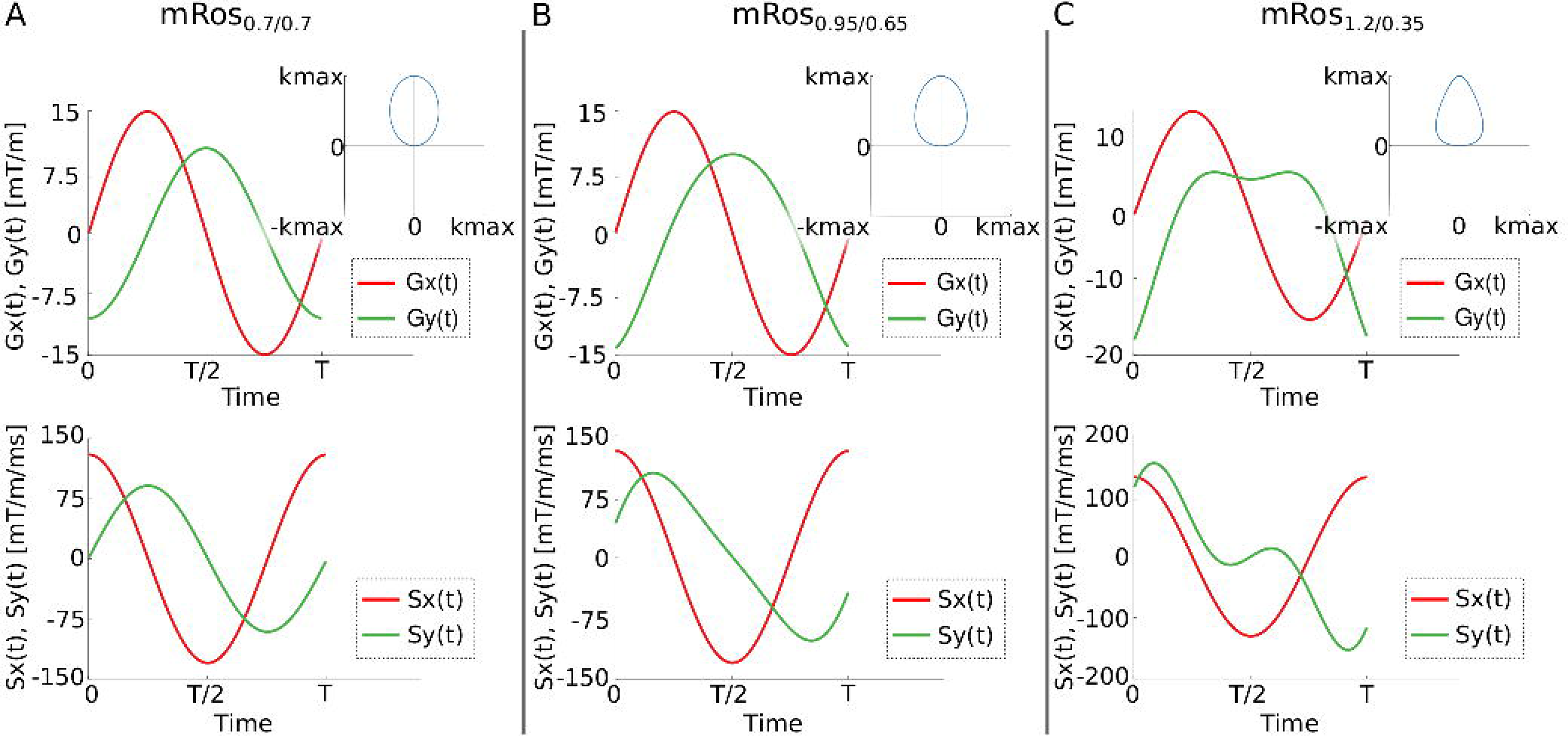
K-space trajectory, gradient amplitudes and slew rate of the modified rosette trajectory for a constant (A) and two sets of time-dependent compression factors (B, C). The gradient moment and slew rate are shown for both phase encoding gradients for one period.

With varying boundary compression factors *E*_1_, *E*_2_, and function *f(t)* to transition between them, various petal shapes can be achieved. Using a time-constant compression factor results in elliptical trajectories (Figure 1, A). Figure 1 (B, C) shows an “egg-shaped” trajectory that maximizes SNR while staying within the hardware limitations.

### 3.2 K-space density

Using the two boundary compression factors, the oversampling of the rosette trajectory in the k-space center and k-space periphery can be reduced. Figure 2 shows the k-space density functions of the rosette and modified rosette trajectories for a one-dimensional line going through the k-space center. Boundary compression factors 1 in the k-space center reduce oversampling since larger circle radii result in a smaller density of sampling points. In the k-space periphery, the desired lower k-space weighting can be achieved using small circle radii. Here a boundary compression factor <1 should be used.

**Figure 2:**
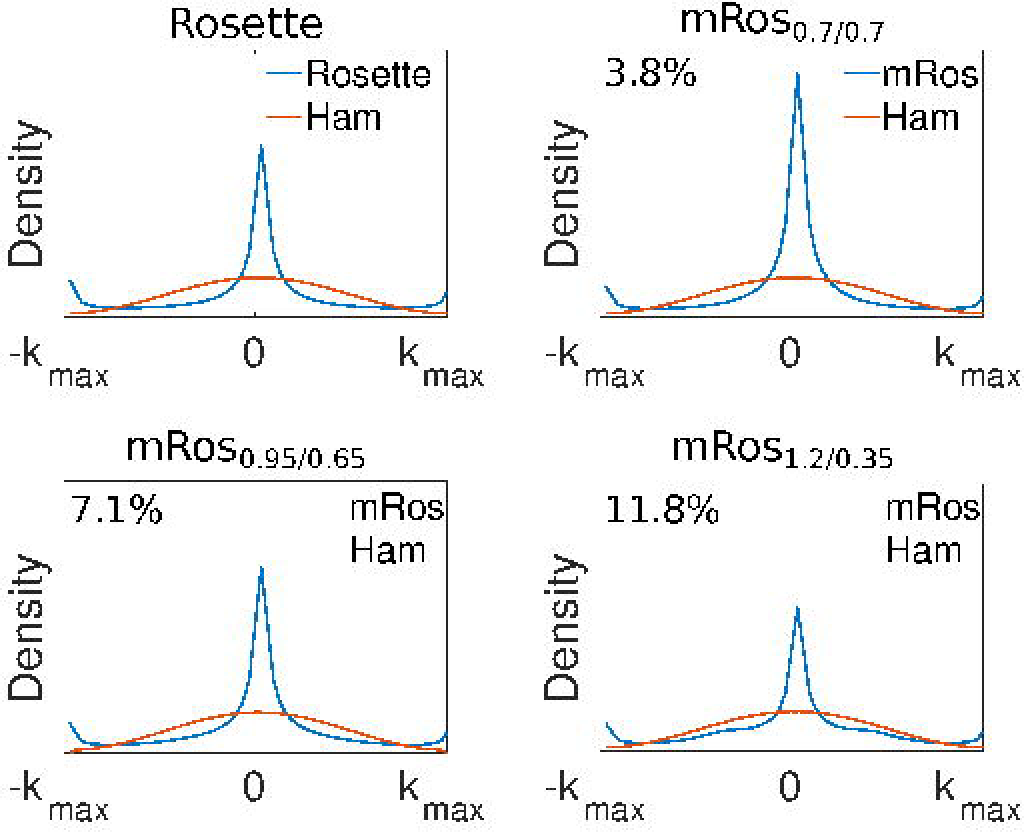
K-space density function of the rosette and modified rosette trajectories for a one-dimensional line with k_y_ = 0 going through the k-space center. The k-space densities of the rosette, mRos_0.7/0.7_, mRos_0.95/0.65_, and mRos_1.2/0.35_ trajectories are shown. The relative SNR of the modified rosette trajectories to the rosette trajectory is written on the top left of each subfigure.

### 3.3 Gradient amplitude and slew rate

The function *e(t)* also influences the gradient amplitude and slew rate. For a given spatial resolution and spectral bandwidth, the gradient amplitude and slew rate are minimized if a constant *e(t)* = 1 is applied. For non-constant *e(t)*, they strongly depend on the difference *E*_1_ − *E*_2_ The boundary compression factors were optimized for highest SNRs with the help of the simulations described in the following sections while staying within the gradient hardware restrictions. For the measurement parameters described later, the resulting trajectories had maximum gradient amplitudes of 14.9 mT/m, 14.9 mT/m, 15.1 mT/m, and 17.8 mT/m for the rosette trajectory, and modified rosette trajectories with three different sets of boundary compression factors (*E*_1/_*E*_2 = 0.7/0.7;_ *E*_1/_*E*_2_ =0.95/0.65; *E*_1/_*E*_2_ =1.2/0.35; referred to as mRos_0.7/0.7_, mRos_0.95/0.65_ and mRos_1.2/0.35_), respectively. The maximum gradient slew rates for the same trajectories were 130.1 mT/ms/m, 130.1 mT/ms/m, 143.9 mT/ms/m, and 191.9 mT/ms/m, respectively. All trajectories were calculated for the same parameters such as spectral bandwidth, number of angular and temporal interleaves, spatial resolution (see point 3.5 for the exact values).

### 3.4 Simulations

Kasper et al. showed that the SNR of a measurement is maximized when the k-space density function matches the desired target function. Thus, when choosing a Hamming filter, the SNR-efficiency of a trajectory can be calculated via the SNR ratio to a hypothetical Hamming filter^25^

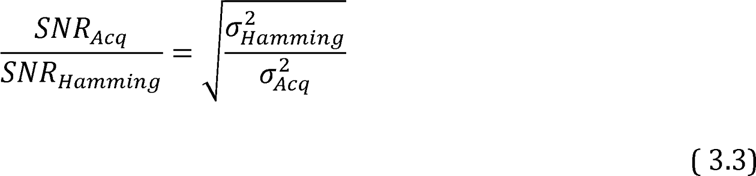

In addition to this analytical approach, the SNR was evaluated using simulated data and applying the pseudo multiple replica-based SNR calculation^27^ (see point 3.6 for more information). Here, the percentage deviation of the SNR of the modified rosette trajectories to the rosette trajectory was calculated, as well as to phase encoding and EPSI with ramp sampling.

### 3.5 Experiments

Data were acquired on a 7T MAGNETOM+ (Siemens Healthineers, Erlangen, Germany) with a 32-channel RF head coil for receive, and a volume coil for receive and transmit (Nova Medical, Wilmington, MA) from a silicon oil and resolution phantom and five healthy volunteers. Written informed consent was obtained from all volunteers.

In addition to the modified rosette trajectories with three different sets of boundary compression factors (mRos_0.7/0.7_, mRos_0.95/0.65_ and mRos_1.2/0.35_), the rosette trajectory was measured. The following parameters were used for both the phantom and in vivo measurements for all trajectories: matrix size 64×64, FOV 220×220 mm^2^, slice thickness 10 mm, resolution 3.4×3.4×10mm^3^, two temporal interleaves, 840 FID points, spectral bandwidth 2778 Hz, ADC bandwidth 666.666 kHz (considering oversampling), and oversampling factor 2. All of the compared trajectories share the same trajectory parameters (i.e., 101 petals, 72 gradient points per petal, 240 ADC points per petal, k_max_ 0.145 mm^-1^, spectral bandwidth 2778 Hz). Two in vivo scans were obtained, one with WET water suppression, four averages and repetition time (TR) 440 ms, acquisition time (TA) 6:04 min:s, and the other without water suppression, one average and TA 1:31 min:s for estimating the coil combination weights. The latter settings were also used for the phantom measurements.

The four trajectories were estimated based on oil phantom measurements using an adapted method of Brodsky et al.^28^. We used the same parameters as for the in-vivo measurements, except: slice thickness 1 mm, slice offsets of 20 mm, TE 6.02 ms, and using the volume coil for signal reception.

A 3D-T_1_-weighted magnetization prepared rapid gradient echo (MP2RAGE)^29^ sequence was measured in the in vivo scan protocol for anatomical reference with a nominal resolution of 1.1×1.1×1.2 mm^3^ and TA of 4:57 min:s. 3D FID-MRSI scans using the mRos_1.2/0.35_ were measured, once with water suppression, and once without. The following parameters were used: matrix size 64×64×17, FOV 220×220×76 mm^3^, two temporal interleaves, 840 FID points, spectral bandwidth 2778 Hz, one average, TR 440 ms, and TA 18:44 min:s. Note that the acquisition of the coil combination weights can be highly accelerated, but this was not the purpose of this paper. Before the measurements, manual shimming was performed. MRSI data were density compensated, filtered with a Hamming filter in k-space, and reconstructed with a type 2 non-uniform discrete Fourier transform using the measured trajectories. A lipid removal using an L1-regularization was performed^30^.

### 3.6 Evaluation

In vivo spectra were fitted using LCModel^31^. Our basis set included 17 simulated brain metabolites and a measured macromolecular background^32^. SNR ratios were calculated for all volunteers and trajectories using the pseudo replica method. The noise was estimated for each voxel using the standard deviation through a stack of pseudo replicas of the true image with added, uniformly distributed, and correctly scaled noise. This is a simulation-equivalent of measuring multiple times the same data, and was shown to provide good estimate for the standard deviation of the noise^25^. The signal was defined as the amplitude of the fitted NAA peak^33^. A mean over all voxels inside a brain mask derived from the T_1_-weighted images was calculated. The mean and standard error over all volunteers is reported. The same comparison is performed for the Cramer-Rao Lower Bounds (CRLB) values of total *N*-acetylaspartate (tNAA), total creatine (tCr), total choline (tCho), glutamate and glutamine (Glx) and *myo*-inositol (*myo*-Ins), the spectral linewidth, and the lipid SNR. The CRLBs and linewidths are calculated by LCModel, while the lipid SNR was calculated by summing the magnitude spectra in the range of 0.0 to 1.8 ppm and dividing by the standard deviation of the noise. Metabolic maps and spectra are shown for the first volunteer in agreement with minimum reporting standards^34^.

Two-sided paired *t*-tests were performed to test for statistically significant differences between SNR, linewidth, CRLB and lipid SNR values of the different modified rosette trajectories to the rosette trajectory.

## 4. Results

### 4.1 SNR Simulations

The k-space density simulations evaluate to an SNR increases of 3.8%, 7.1% and 11.8% to the rosette trajectory for the mRos_0.7/0.7_, mRos_0.95/0.65_, and mRos_1.2/0.35_ trajectories, respectively. Compared to phase encoding the simulations showed significant SNR improvements of 16.6%, 20.19%, 24.5% and 32.01%, 36.10%, 42.08% compared to EPSI for the mRos_0.7/0.7_, mRos_0.95/0.65_, and mRos_1.2/0.35_ trajectories, respectively. Using the pseudo multiple replica method on simulated data, percentage increases compared to the rosette trajectory of 2.0%, 2.6% and 8.7% for the mRos_0.7/0.7_, mRos_0.95/0.65_ and mRos_1.2/0.35,_ respectively, were estimated.

### 4.2 Phantom results

In Supporting Table 1 the calculated SNRs from phantom measurements are reported. For better comparison, the percentage deviation of the mean SNR values calculated over the entire phantom for the modified rosette trajectories to the rosette trajectory is shown. All three modified rosette trajectories showed significantly higher SNRs compared to the rosette trajectory. Over all three measurements the SNR increased in comparison to rosette trajectory by 3.81±0.2% (*p*≪0.001), 7.00 ±0.17% (*p*≪0.001) and 12.47±0.29% (*p*≪0.001) for the mRos_0.7/0.7_, mRos_0.95/0.65_, and mRos_1.2/0.35_ trajectories, respectively. In Figure 3 the reconstructed images from the phantom measurements are shown. No differences can be seen in the images reconstructed with the measured trajectories (Figure 3, bottom row). Using an analytically determined trajectory lead to ringing artifacts and localization errors near the edges of the phantom when the modified rosette trajectory was used (Figure 3, top row). In Supporting Figure 1 the results of the resolution phantom measurement are shown. Similar resolution properties can be observed for the rosette and modified rosette trajectories.

**Figure 3:**
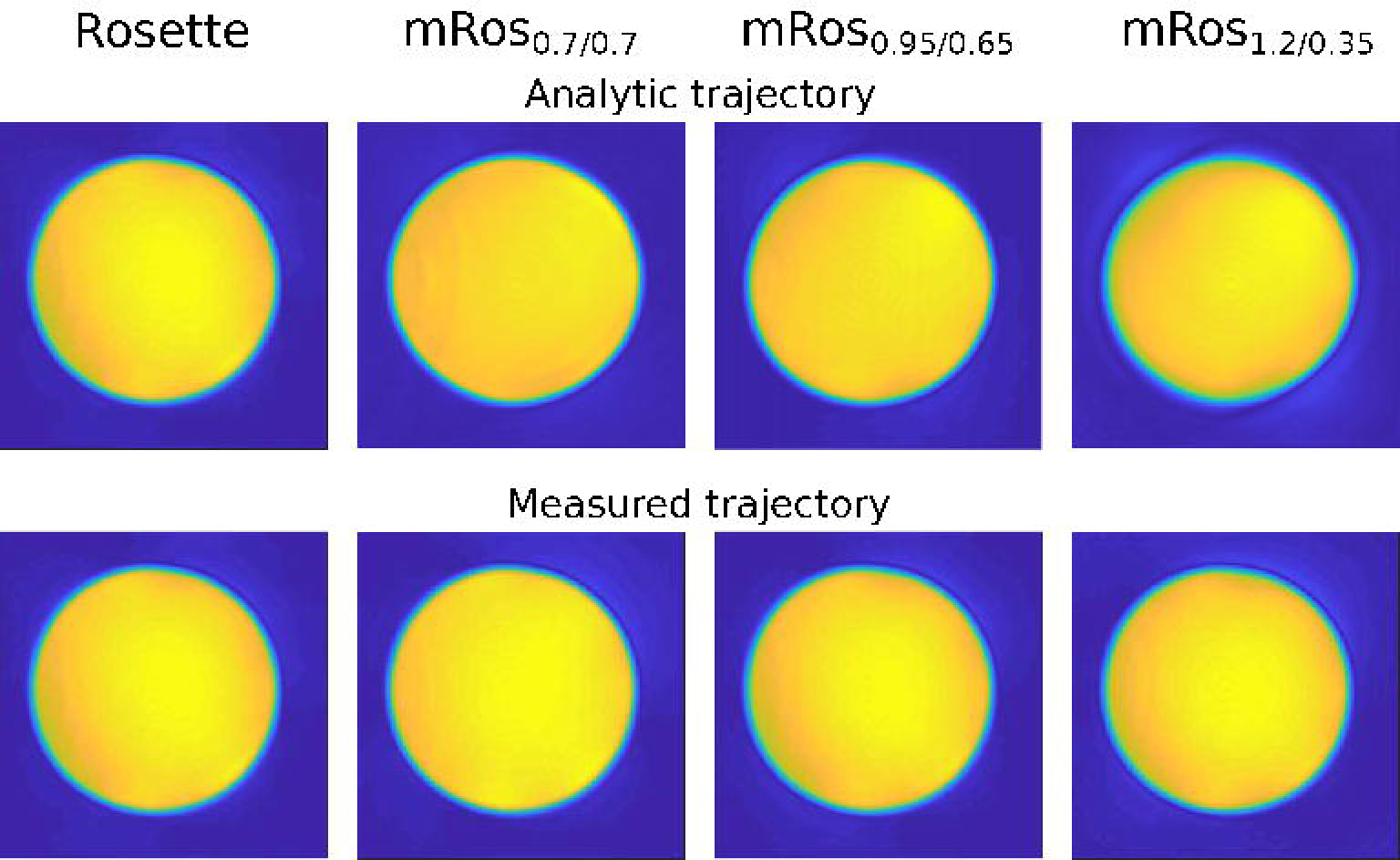
Results from the phantom measurement using a silicon oil phantom and the in vivo protocol (FOV of 220×220 mm^2^, slice thickness 10 mm and matrix size of 64 x 64). The phantom was measured with the rosette, mRos_0.7/0.7_, mRos_0.95/0.65_ and mRos_1.2/0.35_ trajectories. The phantom results were Hamming weighted and zero-filled to obtain a matrix size of 300 x 300. Images were reconstructed with an analytically determined (top row) and measured (bottom row) trajectories. Using a measured trajectory eliminated significant artifacts from gradient trajectory errors that can be observed for mRos_1.2/0.35_.

### 4.3 In vivo results

Supporting Table 2 shows the calculated SNR deviation of the modified rosette trajectories in comparison to the rosette trajectory. Mean SNR gains of 3.0% (*p*=0.015), 6.3±1.1% (*p*≪0.001), and 8.9±1.6% (*p*≪0.001) can be observed over all subjects for the mRos_0.7/0.7_, mRos_0.95/0.65_ and mRos_1.2/0.35_ trajectories, respectively. Representative spectra for the first volunteer at a voxel location specified by the minimum reporting standards^34^ for all compared trajectories are shown in Figure 4. The raw spectra and fitted data are displayed. Good fitting results with similar noise can be observed in the spectra for all trajectories. Metabolic maps for the first volunteer and the major metabolites tNAA, tCr, tCho, and Glx are shown in Figure 5 for 2D and Supporting Figure 2 for 3D data. Hardly any differences between the trajectories are visible. Lipid artifacts can be observed in the 3D tNAA maps. 2D SNR maps for the first volunteer are shown in Supporting Figure 3.

**Figure 4:**
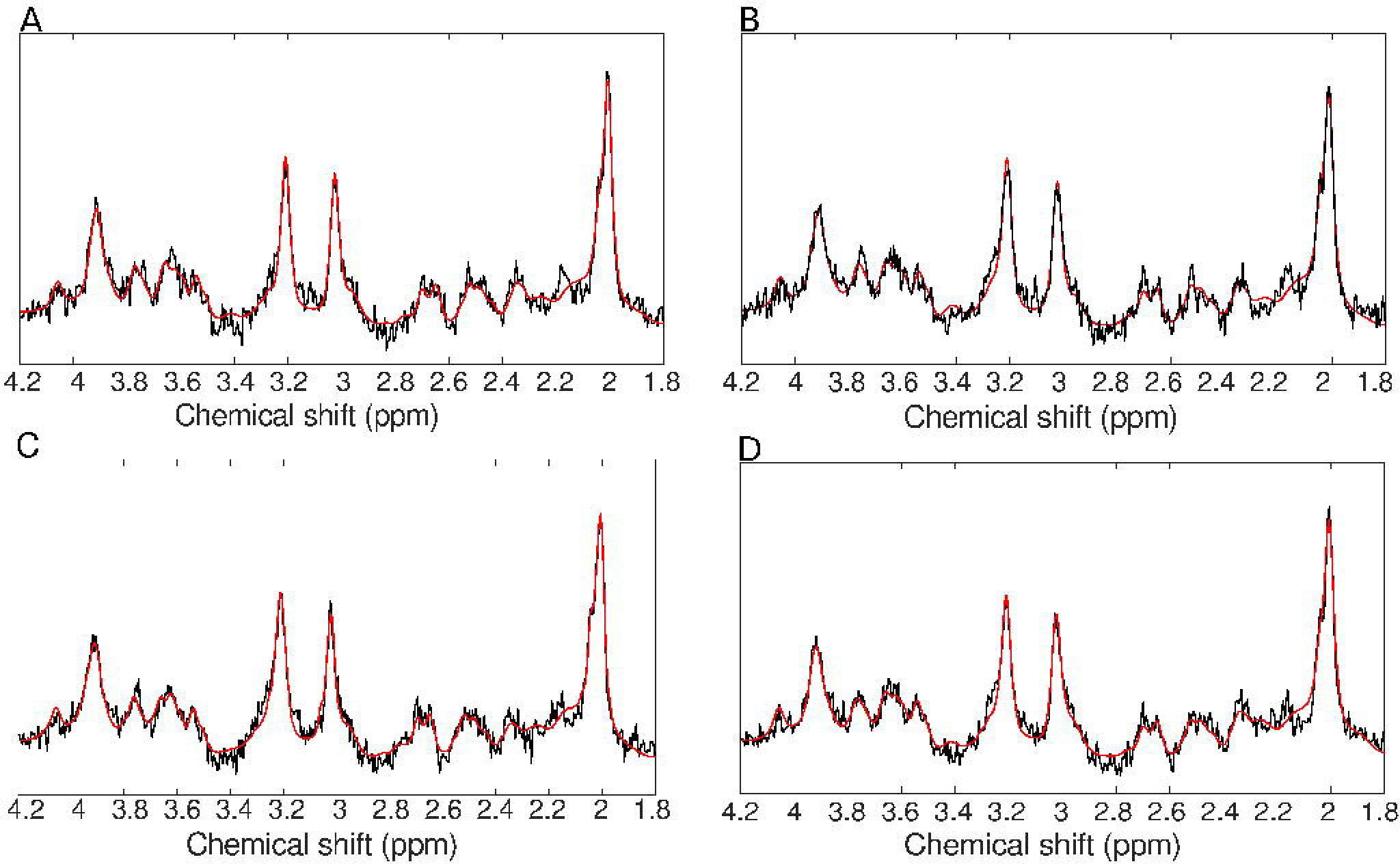
In vivo spectra from the first volunteer and all measured trajectories (rosette (A), mRos_0.7/0.7_ (B), mRos_0.95/0.65_ (C) and mRos_1.2/0.35_ (D)). Fitted spectra (red) and measured data (black) were taken from LCModel. First order phase correction was performed. Similar spectral noise can be observed for the spectra acquired with the rosette, mRos_0.7/0.7_, mRos_0.95/0.65_ and mRos_1.2/0.35_ trajectories. The voxel location from which the spectra are taken is shown in Figure 5.

**Figure 5:**
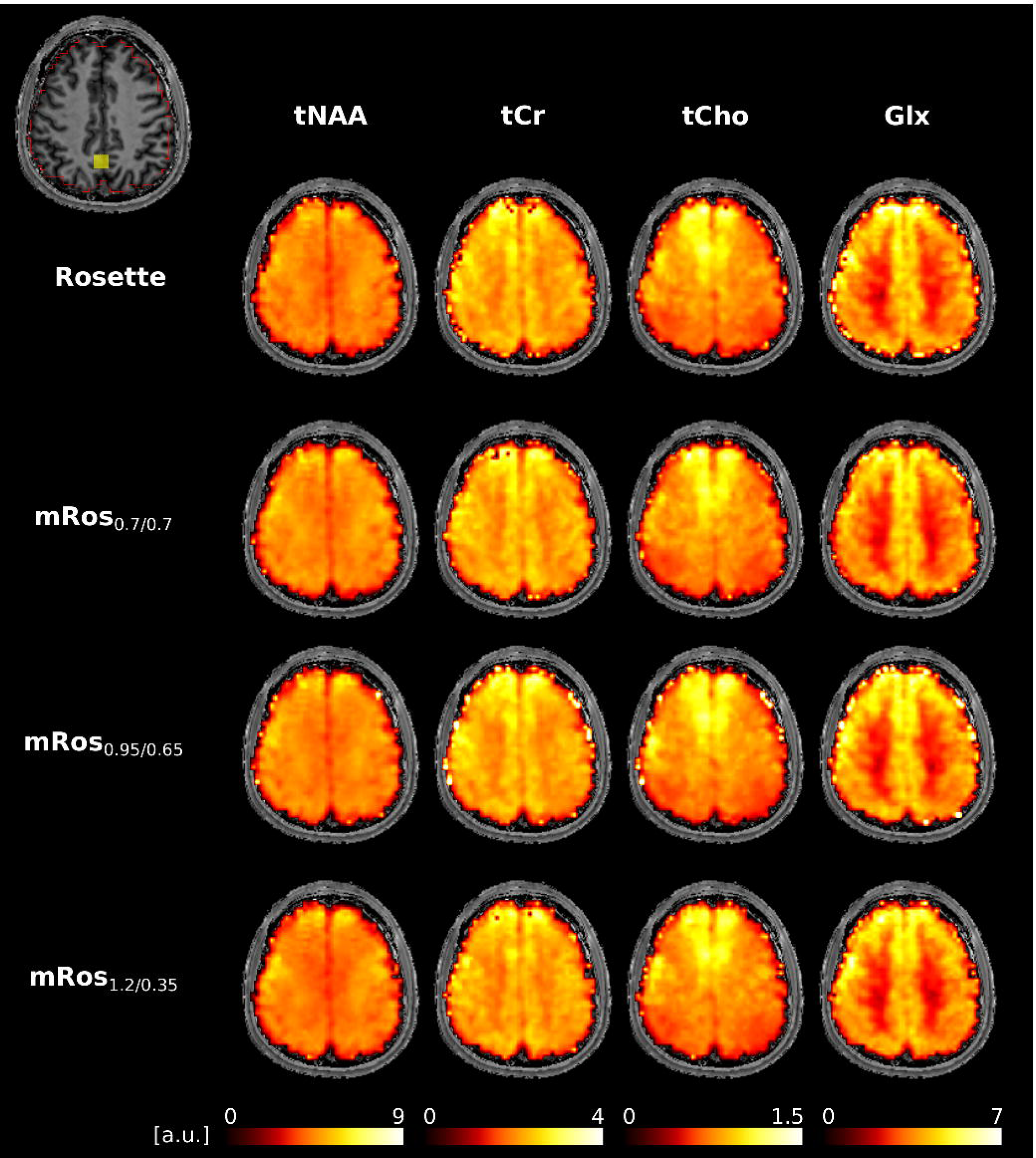
Results from the first volunteer using the rosette, mRos_0.7/0.7_, mRos_1.2/0.35_ and mRos_0.95/0.65_ trajectory (FOV of 220×220 mm^2^, slice thickness 10 mm and matrix size of 64 x 64). Metabolic maps for total *N*-acetyl-aspartate (tNAA), total creatine (tCr), total choline (tCho) and glutamine + glutamate (Glx) are displayed. The ROI and voxel location of the spectra shown in Figure 4 is shown in the top left.

The mean CRLB values for tNAA, tCr, tCho, Glx and *myo*-Ins are shown in Supporting Table 3. All voxels inside the brain mask (Figure 5) had CRLBs<50%. A small non-significant reduction of the mean CRLB values can be observed for all modified rosette trajectories compared to the rosette trajectory. A small increase of the number of voxels with CRLB values lower than 10% can be observed for the mRos_1.2/0.35_ compared to the rosette trajectory. The linewidths were 14.24±0.) Hz, 14.25±0.) Hz, 14.27±0.1 Hz,

14.38±0.1 Hz for the rosette, mRos_0.7/0.7_, mRos_0.95/0.65_, mRos_1.2/0.35_ trajectories, respectively. The lipid SNR normalized to the rosette values were 1.00±0.10, 0.79±0.036, 1.16±0.05, 1.07±0.05. Paired *t*-tests between the values of the modified rosette trajectories and the rosette showed that the CRLB, linewidth and lipid SNR are non-significantly different.

## 5. Discussion

We have shown that by modifying the rosette trajectory higher SNR values of up to +12% can be achieved. The SNR results from phantom and in vivo measurements were consistent with simulations, but a smaller SNR increase was observed in the in vivo data using the mRos_1.2/0.35_ trajectory (Supporting Figure 4). The reason for that might be that this trajectory is more susceptible to subject related artifacts such as motion. The highest SNR was achieved with the mRos_1.2/0.35_ (∼9% higher than rosette), followed by the mRos_0.95/0.65_ (∼6% higher than rosette) and mRos_0.7/0.7_ (∼3% higher than rosette), while the lowest SNR was achieved with rosette trajectory. Because of the similar design of the trajectories and the same number of sampling points along the trajectory, no additional measurement time is necessary to realize this SNR gain. All compared trajectories had similar linewidths, lipid contaminations, and resolution properties. The small and non-significant decrease of the CRLB values for the modified rosette data in comparison to the rosette data indicate that the SNR might lead to a small improvement in metabolite quantification precision.

Kasper et al. showed that the sampling SNR-efficiency is highest, when the measured k-space density matches the desired k-space density^25^. Our desired k-space density was a Hamming filter, which is reasonably well approximated by modified rosette trajectories, and less so by rosette trajectory. Both, the rosette and modified rosette trajectories have mild oversampling of the k-space center. This comes at the disadvantage of less efficient k-space sampling and a greater deviation from the ideal k-space weighting. However, mild oversampling of the k-space center makes the trajectories more robust against subject motion as shown by Schirda et al.^14^.

The slew rate of the mRos_1.2/0.35_ trajectory of 191.9 mT/ms/m was close to the hardware limit of 200 mT/ms/m and noticeably higher compared to that of the rosette and mRos_0.7/0.7_ trajectories, which used 130.1 mT/ms/m. Measurements with the mRos_0.95/0.65_ trajectory resulted in a ∼6% SNR increase. With maximum slew rates of 143.9 mT/ms/m, which are slightly higher than those of the rosette, the mRos_0.95/0.65_ trajectory can be a good compromise between high SNR and reduced gradient stress. A major advantage of modified rosette trajectories is the flexibility to adjust the two compression factors so that a given slew rate restriction is met, while maximizing the SNR. By doing so the sequence can be adapted for several different modalities, such as different magnetic field strengths or matrix sizes. The smallest achievable slew rate is that of the rosette trajectory. All compared trajectories provided similar spectra and metabolic maps, indicating comparable spectral qualities between all trajectories.

In comparison to other SSE trajectories, modified rosette trajectories share the advantages and disadvantages of the rosette trajectory, except for their higher SNR-efficiency. In contrast to spiral and EPSI trajectories, modified rosette trajectories benefit from their self-rewinding properties, which provides higher SNR due to not needing a rewinder gradient. Moreover, the k-space density of modified rosette trajectories is closer to a Hamming filter than most other MRSI trajectories, except the ideally shaped density-weighted CRT sampling^18^. Modified rosette trajectories achieve their k-space density not by measuring additional angular interleaves as is the case for density-weighted CRT, but by changing the gradient shape. Therefore, the modified rosette trajectories do not prolong the measurement while achieving a comparable SNR-efficiency as density-weighted CRT encoding. In addition, the modified rosette trajectories, as well as the rosette use smaller circles than CRT, and therefore can cover much higher spectral bandwidths for a given resolution, which is essential for MRSI at ≥7 T^14^. Modified rosette trajectories share the benefit of circular k-space coverage of other non-Cartesian SSE strategies such as spirals, rosettes and CRT. This results in a more time-efficient k-space acquisition for a given resolution, as including the corners of k-space (e.g., like in EPSI) cause an anisotropic spatial resolution^35^. Another advantage of modified rosette trajectories is that they pass through the k-space center in each circumnavigation, which opens the possibility to correct for instabilities such as motion, frequency drifts or gradient delays without the need for additional measurements^36^.

### Limitations

The gradient amplitudes and slew rates correlate with the compression factors of the modified rosette trajectories. At ultra-high fields B_O_ >7 T this can lead to increased gradient hardware stress, which has to be well balanced. Especially beyond 10 T, where higher spectral bandwidths and spatial resolution are expected, this limits the possible SNR gain. Similarly, higher matrix sizes lead to increased slew rates. This could be mitigated by the use of temporal interleaves, which would, however, increase scan times and could—for ^1^H MRSI—potentially introduce unwanted spectral artifacts associated with unsuppressed water signals. In general, phantom measurements showed a higher susceptibility of modified rosette trajectories to gradient trajectory errors, if no compensation was used, leading to incorrect signal allocation in the reconstructed image^37^. However, this problem was fully corrected for by using measured trajectories. Lipid artifacts were visible in the 3D tNAA maps that were not observed in the 2D images. These might result from an insufficient lipid removal due to increased contamination in the 3D maps. By optimizing the lipid regularization parameter lipid artifacts could be further reduced.

In general, modified rosette and rosette trajectories sample k-space slower than SSE approaches like spirals, CRTs and EPSI. However, at high-resolution, high-bandwidth 3D coverage, these alternatives are not feasible due to gradient performance limitations. Shen et al.^22,23^ showed that using 3D modified rosette trajectories together with CS only 80% of all rosette petals are needed for artifact free images. The possibility of accelerating modified rosette trajectories using CS reconstruction approaches proved useful to a variety of application in recent publications ^38–44^. Modified rosette trajectories will likely require a combination with complimentary acceleration methods such as parallel imaging or compressed sensing to reach clinically attractive scan times.

## 6. Conclusion

We have shown that high-resolution metabolic mapping using modified rosette trajectories benefits from increased SNR efficiency, while staying within the gradient performance limits in comparison to other SSE approaches. Using the two compression parameters, trajectories can be adapted to a variety of applications. This makes rosettes especially interesting for MRSI at ultra-high fields.

## Data Availability

All data produced in the present study are available upon reasonable request to the authors.

## 7. Acknowledgments

### 7.1 Author contributions

This study was funded by Austrian Science Fund (FWF) I 6037-1, FWF P 34198, NIH grant R01EB031787, and National Cancer Institute/National Institutes of Health (NCI/NIH) R01CA255479. MM acknowledges the support of the NIH grants: P41 EB027061 and P30 NS076408.

### 7.2 Financial disclosure

None reported.

## 8. Supporting information

The following supporting information is available as part of the online article:

## Figures

**Supporting Figure 1:** Resolution phantom experiments with the in vivo protocol. Zero-filling to a matrix size of 300×300 was applied for better visualization. Pixel intensities along a horizontal line in the center of the image are shown below. Comparable resolution capabilities can be observed for all compared trajectories.

**Supporting Figure 2:** Whole-brain metabolic maps acquired with the mRos_1.2/0.35_. A matrix size of 64×64×17 was measured in 19 minutes.

**Supporting Figure 3:** SNR maps from the first volunteer for the rosette and modified rosette trajectories.

**Supporting Figure 4:** Boxplot showing the increase in SNR for the modified rosette trajectories compared to the rosette. The results of the phantom and in vivo measurements are shown.

**Supporting Figure 5:** Distribution of petals in k-space of the sequences compared (top row). In the bottom row the analytic and measured k-space trajectory can be observed for the petal with rotation angle a = n (red box in the top row).

