## Supporting Material for "Proton Free Induction Decay MRSI at 7T in the Human Brain Using an Egg-Shaped Modified Rosette K-Space Trajectory"

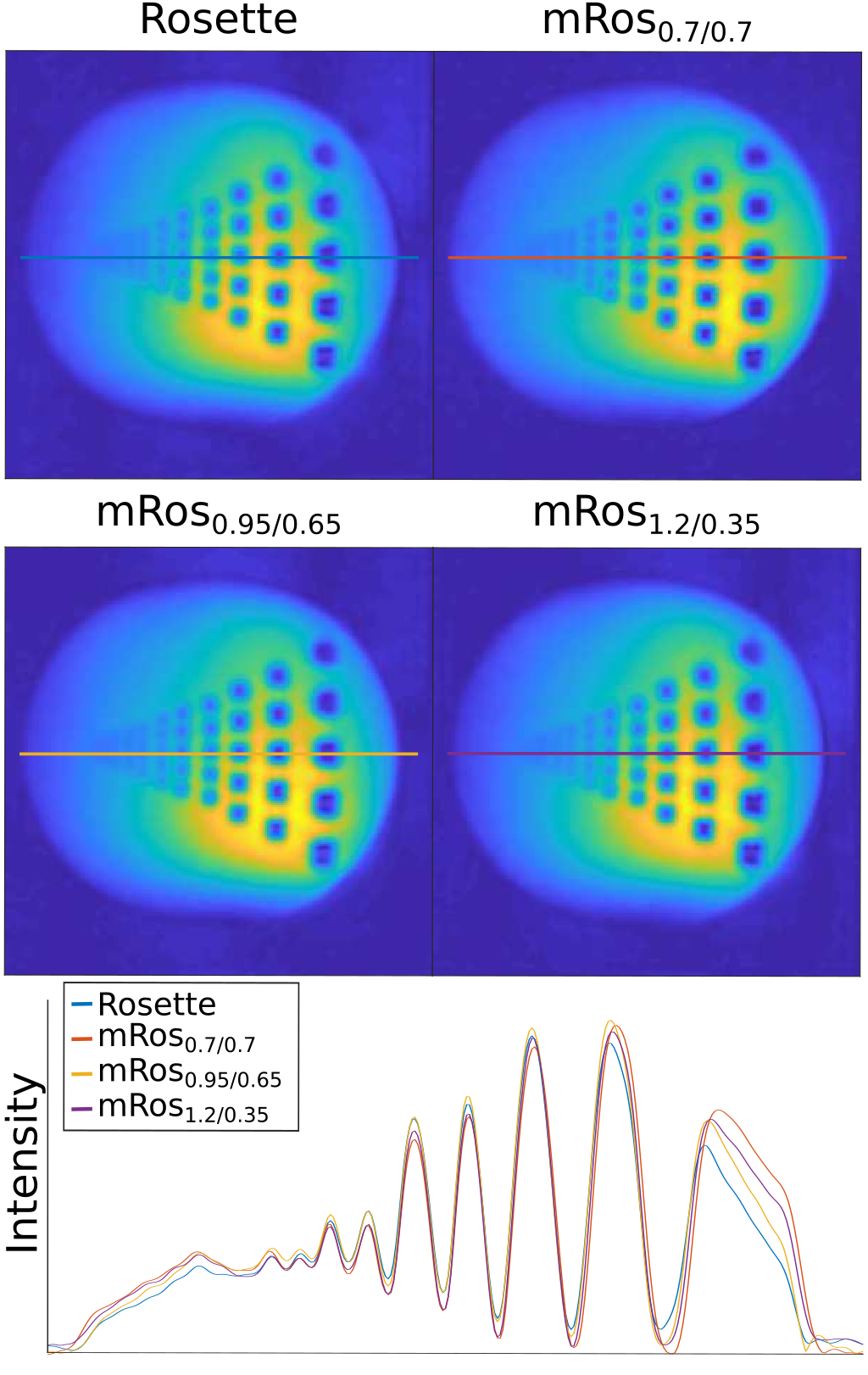

**Supporting Figure 1:** Resolution phantom experiments using the in vivo protocol. Zero-filling to a matrix size of 300x300 was applied for better visualization. Pixel intensities along a horizontal line in the center of the image are shown below. Comparable resolution capabilities can be observed for all compared trajectories.

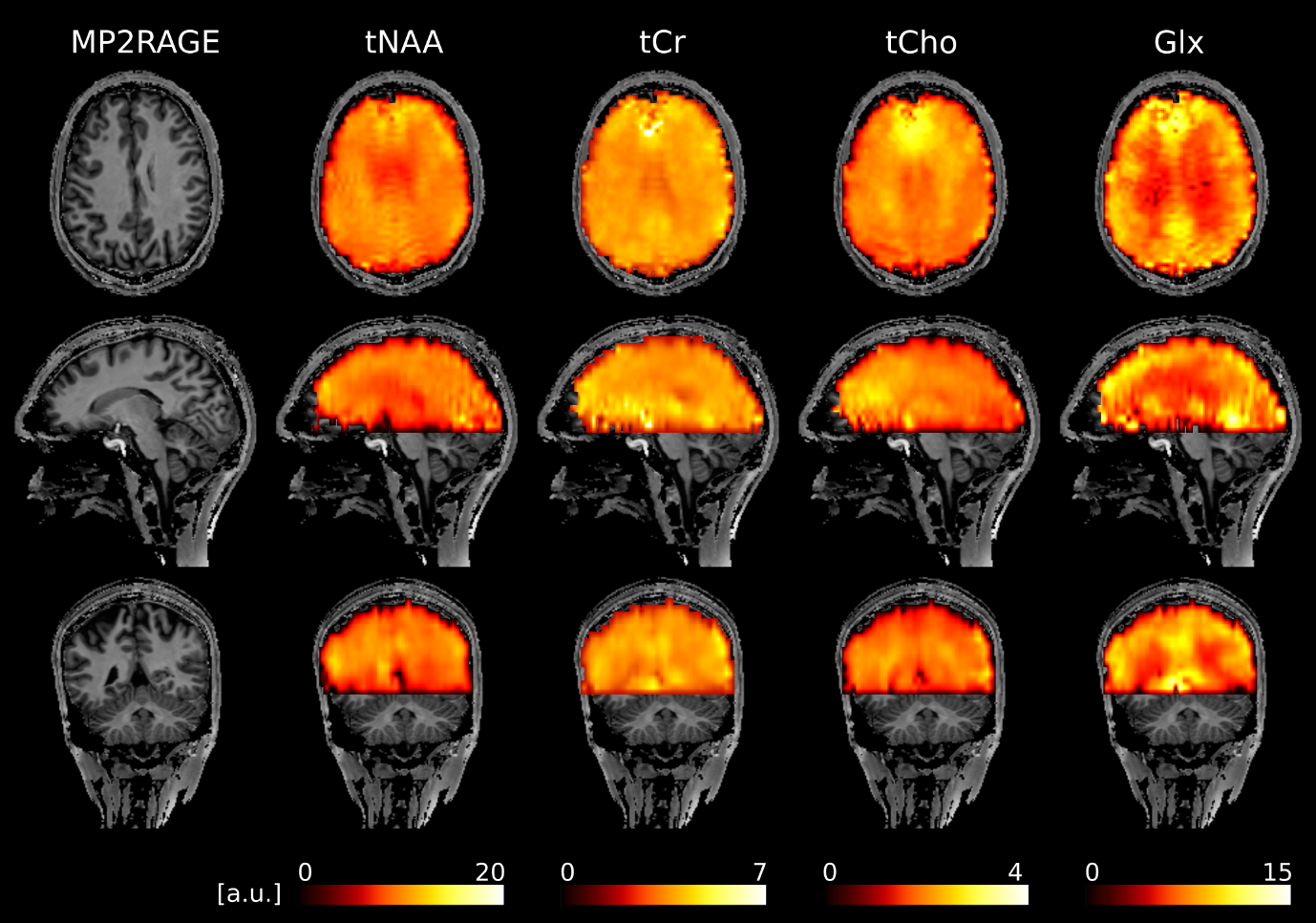

**Supporting Figure 2:** Whole-brain metabolic maps acquired with the mRos_1.2/0.35_. A matrix size of 64x64x17 was measured in 19 minutes.

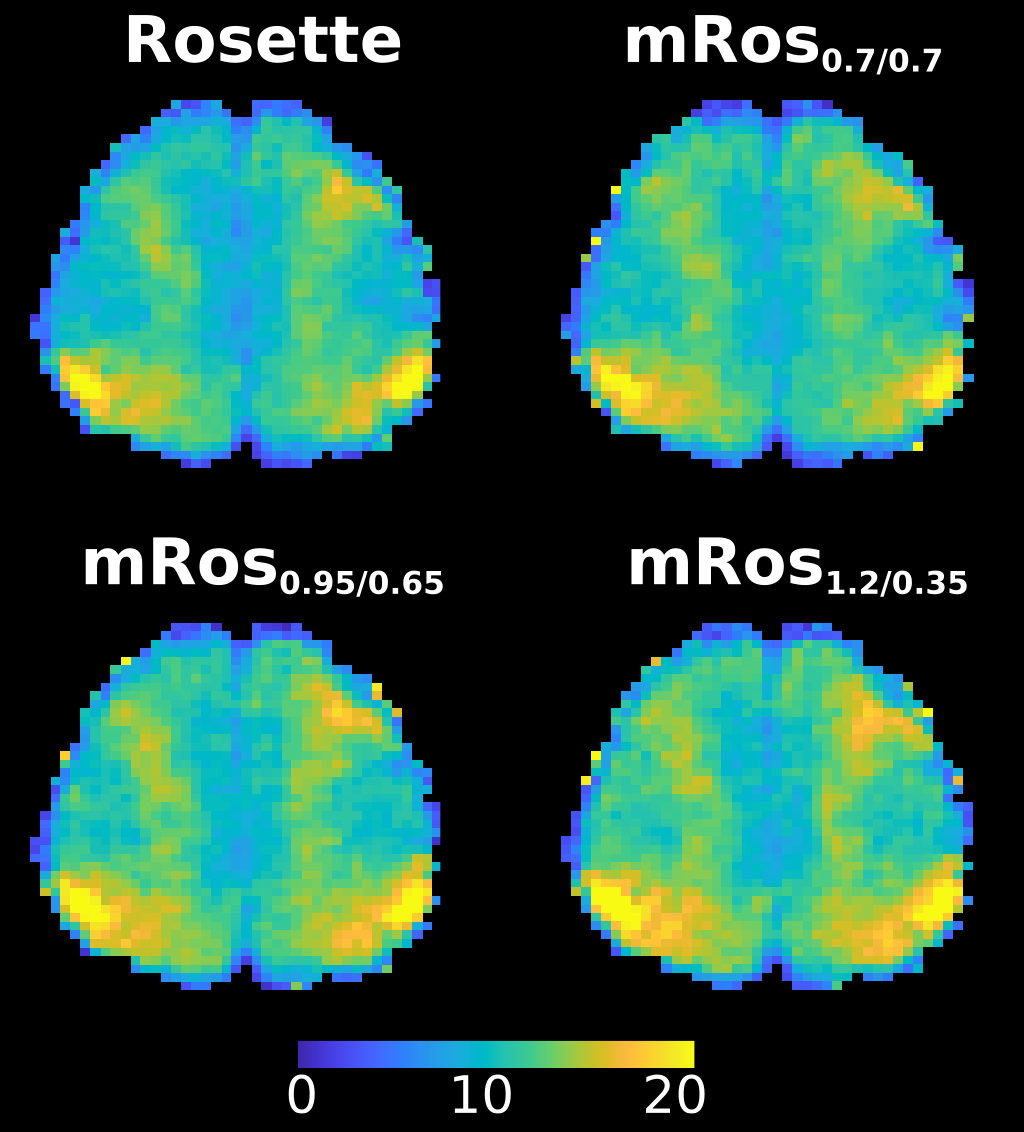

**Supporting Figure 3:** SNR maps from the first volunteer for the rosette and modified rosette trajectories.

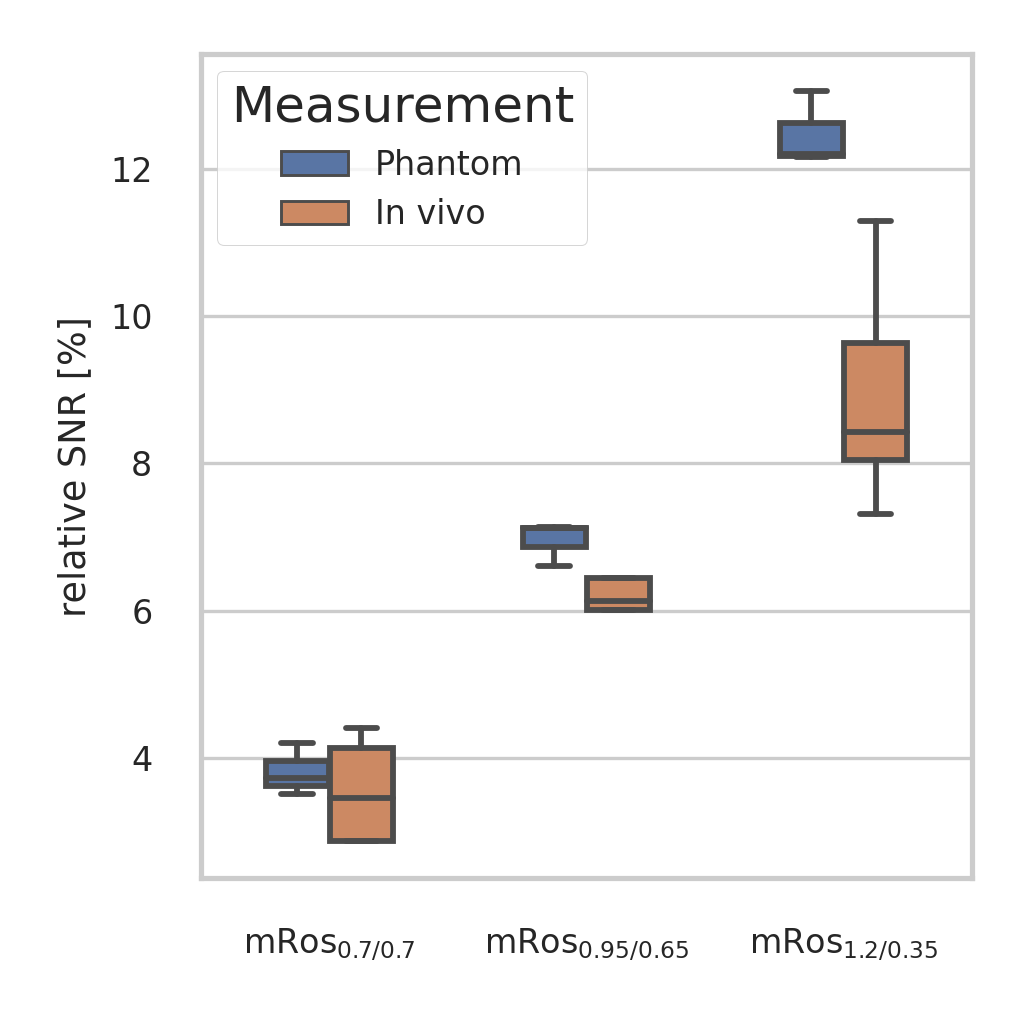

**Supporting Figure 4:** Boxplot showing the increase in SNR for the modified rosette trajectories compared to the rosette trajectory. The results for the phantom and in vivo measurements are shown.

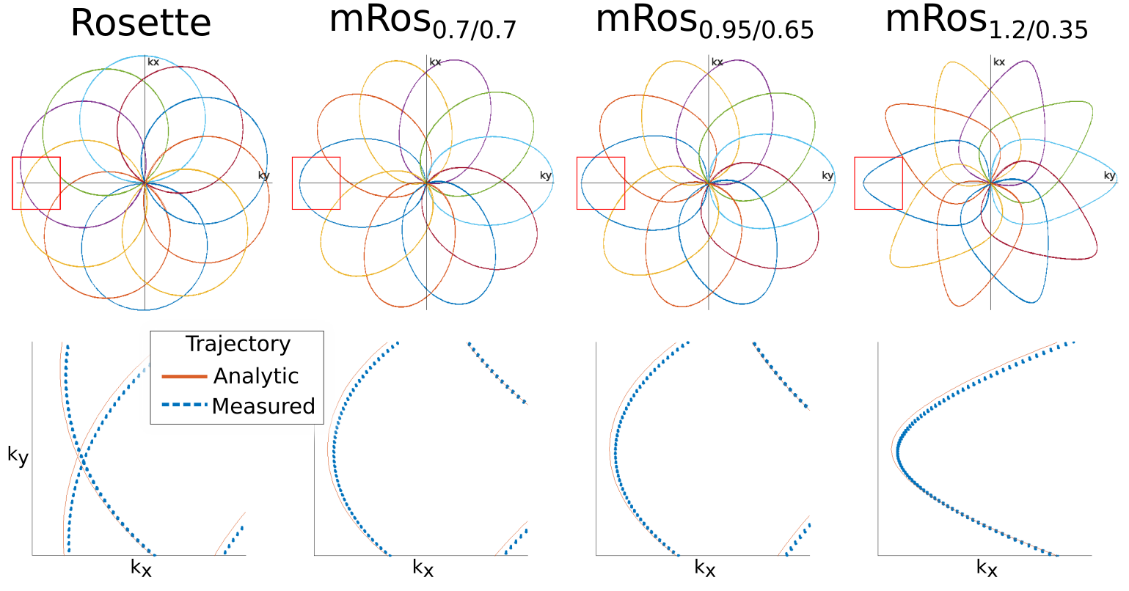

**Supporting Figure 5:** Distribution of petals in k-space of the sequences compared (top row). In the bottom row the analytic and measured k-space trajectories can be observed for the petal with rotation angle $\alpha=\pi$ (red box in the top row).

**Supporting Table 1:** Results of the SNR calculation for the rosette trajectory and modified rosette trajectories from the phantom measurement. SNR values are displayed as the percentage difference from the rosette trajectory. The mean and standard error over all three measurements are shown.

|  | **mRos_0.7/0.7_** | **mRos_0.95/0.65_** | **mRos_1.2/0.35_** |
| --- | --- | --- | --- |
| **Meas 1** | 3.51 | 7.14 | 12.20 |
| **Meas 2** | 3.73 | 6.61 | 12.16 |
| **Meas 3** | 4.20 | 7.12 | 13.05 |
| **Mean** | 3.81± 0.20 | 6.96± 0.17 | 12.47± 0.29 |

**Supporting Table 2:** Mean and Standard Error SNR calculated using the pseudo-replica method over all voxels inside an ROI (see Figure 4) for all volunteers and the trajectories with different compression factors.

|  | **Rosette** | **mRos_0.7/0.7_** | **mRos_0.95/0.65_** | **mRos_1.2/0.35_** |
| --- | --- | --- | --- | --- |
| **Vol 1** | 13.08± 0.09 | 13.53± 0.09 | 14.13± 0.09 | 14.56± 0.10 |
| **Vol 2** | 12.94± 0.09 | 13.48± 0.10 | 13.72± 0.10 | 14.19± 0.10 |
| **Vol 3** | 13.35± 0.10 | 13.74± 0.09 | 14.03± 0.10 | 14.43± 0.10 |
| **Vol 4** | 11.48± 0.09 | 12.00± 0.9 | 12.19± 0.09 | 12.45± 0.10 |
| **Vol 5** | 12.46± 0.11 | 12.48± 0.11 | 13.26± 0.11 | 13.37± 0.11 |
| **Mean** | 12.69± 0.04 | 13.09± 0.04 | 13.50± 0.05 | 13.84± 0.05 |

**Supporting Table 3:** Mean and standard error of the CRLBs over all voxels with CRLB <50% for tNAA, tCr, tCho, Glx and *myo*-Ins. Also the mean and standard error of the number of voxels with a CRLB <10% is shown for the same metabolites.

| Metabolite | Rosette | | mRos_0.7/0.7_ | | mRos_0.95/0.65_ | | mRos_1.2/0.35_ | |
| --- | --- | --- | --- | --- | --- | --- | --- | --- |
|  | CRLB [%] | Number of voxels < 10% | CRLB [%] | Number of voxels < 10% | CRLB [%] | Number of voxels < 10% | CRLB [%] | Number of voxels < 10% |
| tNAA | 3.60± 0.01 | 1087.6± 60.47 | 3.38 ± 0.01 | 1092.6± 60.90 | 3.45± 0.01 | 1092.2± 61.22 | 3.54± 0.01 | 1092.6± 60.77 |
| tCr | 4.93± 0.01 | 1081.2± 59.34 | 4.82 ± 0.01 | 1085.2± 60.37 | 4.83± 0.01 | 1085.0± 60.62 | 4.86± 0.01 | 1086.4± 61.05 |
| tCho | 4.75± 0.01 | 1066.0± 63.10 | 4.63 ± 0.01 | 1072.2± 63.21 | 4.64± 0.01 | 1067.6± 65.81 | 4.69± 0.01 | 1074.2± 64.32 |
| Glx | 7.88± 0.04 | 903.6± 41.84 | 7.64 ± 0.04 | 922.2± 38.81 | 7.72± 0.04 | 926.4± 40.49 | 7.78± 0.05 | 920.0± 40.82 |
| *Myo*-Ins | 10.70± 0.05 | 428.0± 48.72 | 10.66 ± 0.05 | 427.2± 48.68 | 10.26± 0.04 | 498.2± 54.77 | 10.17± 0.05 | 520.0± 50.79 |
